# ADHD and intelligence polygenic scores associations with developmental dimensions in children with attention, learning and memory difficulties

**DOI:** 10.1101/2023.12.08.23299712

**Authors:** Andrea M. Santangelo, Olena Ohlei, Silvana Mareva, Diandra Brkic, CALM team, Lars Bertram, Joni Holmes, Duncan Astle, Kate Baker

## Abstract

Common genetic variants make a significant contribution to neurodevelopmental characteristics such as cognitive abilities and ADHD symptoms. The relevance and structure of these associations amongst children with transdiagnostic difficulties in cognition, attention and learning has not been explored. Polygenic scores (PGS) derived from the largest genome-wide association study (GWAS) data at the time of this study on ADHD (38,691 individuals with ADHD and 186,843 controls) and Intelligence (269,867 individuals) were calculated for 524 children and young people (5-18 years old) referred to the Centre for Attention, Learning and Memory (CALM). PGS-trait associations were assessed via linear regression analyses, for a range of cognitive and behavioural dimensional measures, and factor scores from a hierarchical model of psychopathology. PGS associations were explored with and without co-varying for socio-economic status (SES). Within this sample, we found the expected positive associations between ADHD-PGS and ADHD primary symptoms, and between Intelligence-PGS and IQ. ADHD-PGS were also associated with broader externalising behaviours and intelligence scores, and these associations remained significant after removing ADHD-diagnosed participants, or after covarying with SES. Intelligence-PGS showed associations with verbal and non-verbal cognitive skills, but no significant associations with ADHD traits were detected. For the hierarchical model of psychopathology, ADHD-PGS, but not intelligence-PGS, showed associations with the general mental health factor, externalising factor, and social maladjustment factor, only when SES was not included as a covariate. In summary, PGS for neurodevelopmental traits may contribute to both general and specific cognitive and behavioural dimensions in a paediatric transdiagnostic sample. Future studies investigating PGS associations with neural correlates, as well as gene-by-environment interactions, will contribute to our understanding of developmental pathways and risk-resilience mechanisms in child mental health.

## Introduction

Children with neurodevelopmental difficulties often do not meet criteria for a specific diagnosis, or they meet criteria for multiple diagnoses. Whilst case–control designs or population-based studies have been instrumental in establishing disorder-specific and population-level genetic associations, these approaches have shown limited ability to capture the substantial phenotypic and genetic heterogeneity that characterises psychiatric and neurodevelopmental conditions. Individuals with sub-diagnostic or transdiagnostic symptoms are often underrepresented: in case/control studies with samples enriched in extreme phenotypes due to strict inclusion/exclusion criteria; and in general population studies, with samples including unselected individuals, where the genetic effects of sub-diagnostic symptoms may be attenuated due to low prevalence of clinical symptoms. Recognising these complexities, investigations of neurodevelopment and mental health have increasingly adopted a transdiagnostic framework [1]. Within this framework, it is not necessary to meet formal diagnostic criteria to be included in a study, neither is the presence of co-occurring difficulties an exclusion criterion i.e. better mimicking the lived experience of multi-dimensional neurodevelopmental difficulties. Whether the underlying genetic architecture of neurodevelopmental dimensions within transdiagnostic samples of children with heterogeneous and co-occurring difficulties is different from diagnostic samples and general populations is still unknown.

Attention-Deficit/Hyperactivity Disorder (ADHD) is a common neurodevelopmental disorder characterised by symptoms in either the inattentive or hyperactive/impulsive domains, or both [2]. ADHD affects approximately 5% of children and 2.5% of adults, with a high heritability of approximately 70-80% estimated from twin-based studies [3], and high comorbidity with other psychiatric conditions [4]. At the time of this study, the latest genome-wide association meta-analysis (meta-GWAS) of ADHD case-control study identified 27 genome-wide significant risk loci, including genes enriched for expression in early brain development, with 14 % of SNP heritability accounting for ADHD [5].

A proportion of children with symptoms of ADHD or other behavioural difficulties also have cognitive and learning-related difficulties. The ability to think abstractly and problem solve independently of any previously acquired knowledge is known as fluid intelligence [6]. Fluid intelligence is highly heritable, with a general factor of intelligence (*g*) accounting for about 58% of the genetic variance in cognitive traits [7]. A large-scale, meta-GWAS for the *g* factor of general intelligence has identified 205 genomic loci implicating brain-expressed genes involved in nervous system development, neuron differentiation, and synapse structure and activity [8].

Polygenic scores (PGS) are estimates of individual common genetic variation associated with a trait or disorder and can provide insights into the genetic architecture and underlying mechanisms contributing to complex phenotypes [9, 10]. Higher ADHD-PGS have been associated with ADHD diagnosis in independent case-control samples [11], as well as with ADHD symptoms in population-based studies [5, 12, 13], suggesting that an ADHD diagnosis represents the extreme expression of one or more heritable quantitative traits. In addition, ADHD-PGS have also been negatively correlated with cognitive abilities including general intelligence, attention, verbal and non-verbal reasoning, and working memory [5, 14]. Conversely, genetic correlation and Mendelian randomization analysis showed a negative causal association between intelligence-related variants and ADHD and depression symptoms [6, 8].

Shared genetic contributions across disorders are well-recognised [15–17], with ADHD sharing 84–98% of risk variants with other psychiatric disorders [5, 11]. Thus, PGS may influence aspects of neurobiological and cognitive development relevant to a diverse set of educational, behavioural and psychiatric vulnerabilities [7, 18]. Whilst examining specific PGS-behavioural relationships has value and builds on previous literature, children with co-occurring developmental difficulties experience a combination of cognitive, behavioural, social and emotional challenges, that may be better understood in an integrated fashion. Hierarchical models of psychopathology, such as the HiTOP framework [19–21], offer a structured way to examine how genetic liability relates to mental health across multiple levels of phenotypic organization. The HiToP model proposes a p-factor of general psychopathology at the apex, under which more specific factors are described [22, 23]. The p factor is highly heritable (50-60% from twin studies), and the model aligns well with current understanding of the higher order genetic structure of psychopathology. Although ADHD polygenic risk has previously been linked to HiTOP-related factors [12, 24–26], most evidence comes from population-based or adult samples, leaving it unclear whether these hierarchical patterns generalize to clinically referred paediatric populations characterized by high comorbidity and neurodevelopmental heterogeneity.

Following this approach, the Centre for Attention Learning and Memory in Cambridge (CALM, http://calm.mrc-cbu.cam.ac.uk/) recruited a cohort of approximately 800 children recognised by educational or clinical professionals as having difficulties in attention, learning and/or memory [27]. This unique, transdiagnostic paediatric sample provides an opportunity to study the factors (genetics and environmental) and mechanisms that drive diversity in neurodevelopment, independent of categorical diagnoses and their co-occurrence. The CALM cohort is a deep-phenotyped sample from the point of view of behavioural features [23, 28–30], dimensions of cognitive and executive functions [31–33], dimensions of mental health [23, 34], and neuroimaging [35, 36].

A transdiagnostic design enables the investigation of ADHD and intelligence PGS across multiple diagnostic categories and symptom dimensions, thereby allowing the detection of shared, divergent, and moderating genetic effects that may not be observable in diagnostically homogeneous samples. Within a transdiagnostic sample enriched for individuals with learning difficulties, ADHD and intelligence PGS may show associations with cognitive, behavioural, and emotional outcomes that cut across traditional diagnostic boundaries, reflecting pleiotropic genetic influences and dimensional symptom expression. For example, ADHD PGS may relate to attentional or executive functioning deficits in individuals without a formal ADHD diagnosis, while intelligence PGS may exert protective or compensatory effects on cognitive outcomes across diverse psychiatric presentations. Such cross-disorder and moderating effects are likely to be attenuated in population cohorts due to low symptom burden, and cannot be adequately examined in single-disorder case– control studies. Consequently, transdiagnostic analyses provide a more mechanistically informative framework for understanding how polygenic liability for ADHD and intelligence contributes to clinical heterogeneity and functional outcomes across psychiatric disorders.

The current paper addresses the question of whether and how ADHD and Intelligence PGS are associated with relevant neurodevelopmental characteristics within the transdiagnostic CALM sample. We focused on ADHD and Intelligence PGS because these traits reflect important aspects of behavioural and cognitive heterogeneity within the CALM cohort, and there are robust large-scale GWAS studies to draw on for planned analyses. In addition, we conducted association analyses with and without co-variation with socio-economic status (SES), approximated via the Index of Multiple Deprivation (MDI), an important demographic factor that contributes to behavioural symptom likelihood and correlates with polygenic variation [37].

Our first objective was to establish whether ADHD and Intelligence PGS showed associations with their respective primary traits i.e. ADHD symptoms and cognitive test scores, within the CALM sample. Our second objective was to examine whether PGS associations were specific to these traits, or extended to additional behavioural traits (externalising and internalising problems) which would suggest cross-over between behavioural and cognitive dimensions. Thirdly, we explored whether PGS could contribute to our understanding of a hierarchical model of psychopathology (HiToP) within this population. The extent to which PGS show specific or shared associations within the CALM sample could provide insight into the inter-relationships between dimensions of neurodevelopmental difficulty, at a behavioural or mechanistic level.

## Methods

### Participants

A cohort of children aged 5 to 18 years was recruited by the Centre for Attention, Learning and Memory (CALM, http://calm.mrc-cbu.cam.ac.uk/) between February 2014 and December 2021 [27]. The cohort consisted of 800 children referred by a health or educational professional because of difficulties related to attention, learning and/or memory. In addition, 200 children were recruited from the same schools as controls. This paper reports analysis on the referred children only. Comprehensive phenotyping was carried out for all children (for details see Holmes et al., 2019) (**Table 1**), and an optional DNA sample (saliva) was provided. A total of 615 samples were submitted from the referred children for genotyping. After QC filtering (see below), a final N=524 participants remained, with 361 males and 163 females, and average age of 9.5±2.4 years (**Table 2**). Index of Multiple Deprivation (IMD) was used to approximate the socioeconomic status of the sample [38]. Scores for postcode areas in the United Kingdom range from 1st to 32844th (most to least deprived). The range of IMD for the sample indicated participants came from areas with varying degrees of deprivation, with an average ranking above the national median (**Table 2**). The decile distribution for IMD in this cohort is shown in **Fig. S1**. Within this group of participants, 38% had received a diagnosis of any developmental or psychiatric disorder from community services, 22.5% had received a diagnosis of ADHD, 11.8% had been diagnosed with dyslexia or were receiving speech and language therapy, and 7% had received a diagnosis of autism.

**Table 1:**
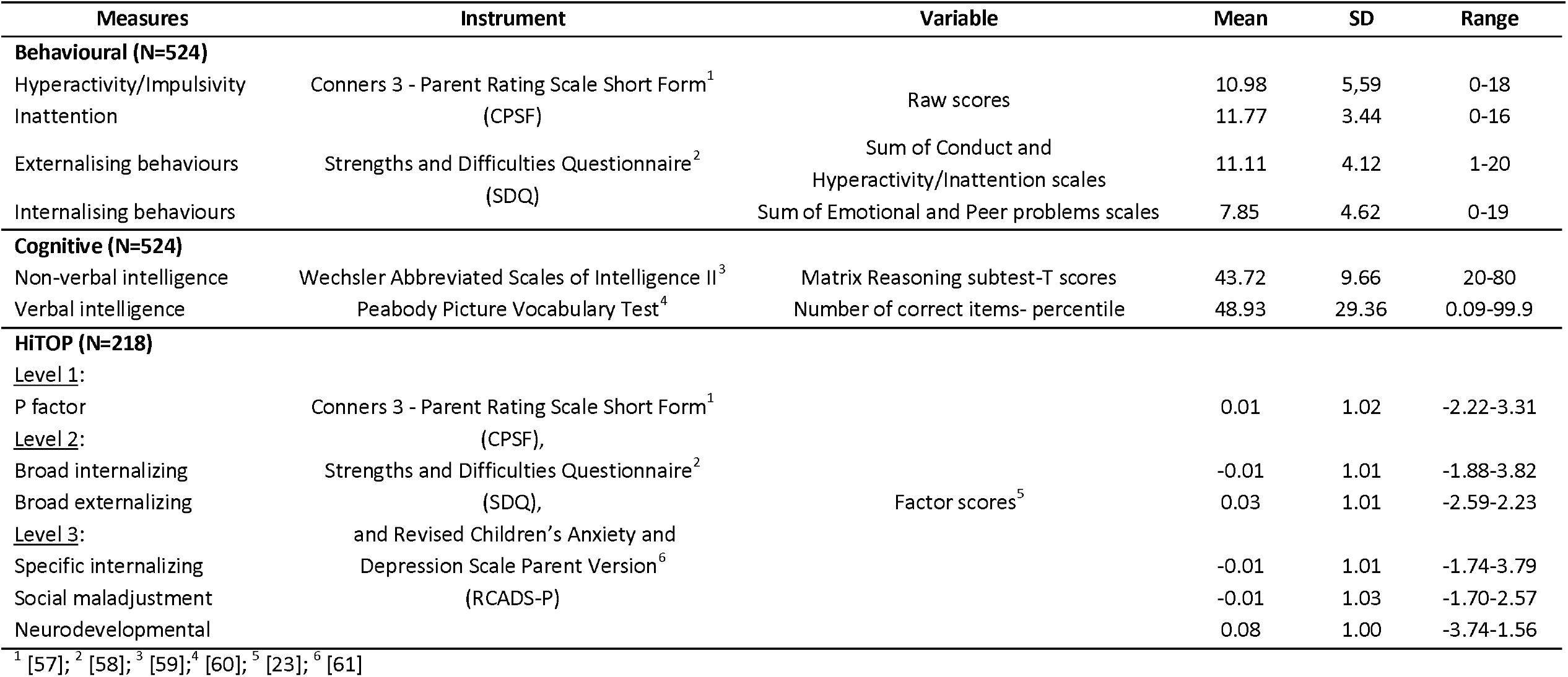
Cognitive and behavioural measures, and HiTOP factor scores, for the CALM sample included in the PGS analyses (after QC filtering)

**Table 2:** Description of the CALM sample after QC filtering.

| **Sample size** | **Total** | **Males/Females** |
| --- | --- | --- |
| N | 524 | 361/163 |
| **Diagnosis** | **Percentage of total** | **N** |
| Any developmental or psychiatric disorder | 38% | 199 |
| ADHD | 22.5% | 118 |
| Dyslexia/speech therapy | 11.8% | 34/28 |
| Autism | 7% | 37 |
|  | **Mean±SD** | **Range** |
| Age (years) | 9.5±2.4 | 5.2-18.6 |
| Index of Multiple Deprivation (IMD) | 20328±8254 | 155-32803 |

### Phenotypic measures

The measures used in the current analysis are listed in **Table 1**, together with descriptive data for each measure used within the CALM PGS-analysed sample.

The Conners 3 – Parent Rating Scale Short Form (CPSF) is a parent/carer-reported questionnaire used to assess symptoms commonly associated with ADHD. It includes 45 items describing behavioural difficulties observed over the past month, which are grouped into six subscales: Inattention, Hyperactivity/Impulsivity, Learning Problems, Executive Function, Aggression, and Peer Relations. We used for our analyses the subscales of Inattention and Hyperactivity/Impulsivity.

The Strengths and Difficulties Questionnaire (SDQ) is a 25-item parent/carer-rated measure that assesses children’s emotional and behavioural functioning over the past six months. It includes five subscales: Emotional Symptoms, Conduct Problems, Hyperactivity/Inattention, Peer Relationship Problems, and Prosocial Behaviour. We used the ‘externalising’ (sum of the conduct and hyperactivity scales) and ‘internalising’ (sum of the emotional and peer problems scales) scores in our study.

The Revised Children’s Anxiety and Depression Scale – Parent Version (RCADS) is a 47-item questionnaire completed by parents/carers to measure the frequency of symptoms related to anxiety and depression. It provides subscale scores for Separation Anxiety Disorder, Social Phobia, Generalized Anxiety Disorder, Panic Disorder, Obsessive–Compulsive Disorder, and Major Depressive Disorder. This questionnaire was only included within the HiToP model (see below).

HiToP factor scores were obtained according to previously published method [23]. Briefly, higher-order dimensions of psychopathology within the CALM cohort were determined using Goldberg’s bass-ackward factor analytic method [39]. This method is based on multivariate hierarchical models of 1 factor, 2 factors, 3 factors and so on, which enables the sequential, top-down extraction of dimensions whilst retaining the shared variance among the measures (the model ceases at the maximal number of factors that can be extracted). All six RCADS subscales were included in the hierarchical dimensional model to comprehensively represent internalizing difficulties. This approach was preferred over the SDQ Emotion subscale, which combines anxiety and depression into a single score. Subscales from the CPSF were then added to capture externalizing symptoms and neurodevelopmental difficulties, specifically Inattention, Hyperactivity/Impulsivity, Executive Function, Aggression, and Peer Relations. CPSF was prioritized over SDQ at this stage due to its broader coverage of neurodevelopmental symptoms. Finally, two SDQ subscales—Conduct Problems and Prosocial Behaviour (reverse-coded to reflect low prosocial behaviour)—were included to supplement the model with domains not captured by the other measures. To avoid bias in the dimensional structure, only one indicator per symptom domain was selected, ensuring balanced representation across all symptom types. See supplementary **Figure S2** for the diagram of the HiTOP model used in this study. Because the RCADS questionnaire was introduced later into the CALM study protocol, HiToP factor analysis was conducted for the subgroup of participants with all measures available, for whom genetic data was available (N=218 individuals after QC filtering).

### DNA extraction, genotyping, QC filtering, imputations, and PGS calculation

For detailed methods, see Supporting Material. In brief, DNA samples were collected from saliva using the Oragene® DNA self-collection kits (DNA Genotek Inc). DNA extraction was performed using prepIT·L2P Kit (DNA Genotek Inc) following manufacturer instructions. Genome-wide SNP genotyping was carried out using the Global Screening Array with shared custom-content (GSA, Illumina, Inc., USA) on an iScan instrument. The raw intensity data was subjected to genotype calling, extensive pre-imputation quality control and SNP imputation using an automated computational workflow described previously, which includes post-imputation step to only retain autosomal SNPs with minimac3 Rsq ≥0.3 (Hong et al., 2020, 2021). A total of 18 samples were excluded (see Supporting Material for details). The LD pruned dataset was used for principal component analysis in PLINK [42] along with the reference dataset of the 1000 Genome Project Consortium Phase 3 (The 1000 Genomes Project Consortium, 2015) to ancestry groups. Only European-descent samples were used in the subsequent analyses, removing 40 non-European samples. Genotype imputation was performed with MiniMac3 [43] software using the “Haplotype Reference Consortium” v1.1 reference panel (McCarthy et al., 2016). Overall, this procedure resulted in 39,131,578 imputed SNPs.

We computed PGS as described previously (Choi et al., 2020). Allele status and effect-size estimates were taken from the summary statistics of the largest ADHD [5] and Intelligence [8] meta-GWAS published to date. Variant QC filtering was performed on both GWAS databases: duplicate and ambiguous SNPs were removed, and only SNPs with MAF >0.05 and imputation quality r^2^ >0.8 were included. The target CALM sample did not overlap with the GWAS datasets. GWAS and CALM genotyping data were annotated on the GRCh37/hg19 genome build. Further 9 samples were removed due to extreme heterozygosity (more than 3 SD from the mean), and 24 closely related individuals were also removed, giving a final sample size of N=524. Variant matching and recoding between the ADHD/Intelligence GWAS base and CALM target datasets was performed using R (version 3.5.3), resulting in 5,583,323 and 3,994,342 overlapping SNPs for ADHD and Intelligence, respectively. After SNP-clumping, PGS were computed for a variety of P-value thresholds (P_t_) in the primary GWAS data (0.001, 0.05, 0.10, 0.20, 0.30, 0.40, 0.50) using PLINK.

### Statistical analysis

PGS-trait association analyses were conducted on standardized values using R (v4.5.0) for each P_t_. ADHD and Intelligence PGS were treated as independent variables in linear regression models adjusting for sex, age, and the first 6 components of a PCA performed on the pruned SNP pool dataset to account for population stratification. We performed sensitivity analysis including index of multiple deprivation (IMD), a proxy for socioeconomic status (SES), as a covariate to reduce potential bias arising from social stratification and gene–environment correlation in polygenic score analyses [37]. As discussed by Abdellaoui and Verweij (2021), polygenic scores for behavioural traits may capture not only direct genetic effects but also indirect genetic and environmental pathways that are correlated with social background. SES is associated with both ADHD polygenic scores and behavioural outcomes, partly because heritable traits related to education and cognition influence family environments and social positioning. Adjusting for SES helps to improves the interpretability of the estimated associations. We also performed sensitivity analysis removing participants that have been diagnosed with ADHD to assess whether significant ADHD-PGS associations were driven by these participants. The variance explained (r^2^) was derived by comparing results from the full model (outcome phenotype with PGS and covariates) vs the null model (outcome phenotype with covariates only). P_t_ values that explained the highest phenotypic variance (max r^2^) were selected. Significant PGS-trait associations were determined after Benjamini-Hochberg adjustment to control for false discovery rate [45], (Tables S1-S5).

We included index of multiple deprivation (IMD) a proxy for socioeconomic status (SES), as a covariate to reduce potential bias arising from social stratification and gene–environment correlation in polygenic score analyses.

As a post-hoc step to aid interpretability, statistical power for ADHD- and Intelligence-PGS was estimated using AVENGEME R package [46] assuming a highly polygenic architecture. SNP-heritability estimates were taken from the discovery GWAS (h^2^ = 0.14 for ADHD; h^2^ = 0.19 for intelligence), with π set to 0.03 and 0.10 respectively. Target sample size was constant across traits (N = 524 for behavioural and cognitive measures, N= 405 after removing ADHD diagnosed participants, and N= 218 for the HiTOP factor analysis). Observed R^2^ values from regression models are reported alongside predicted power (Table S6).

## Results

### ADHD-PGS and Intelligence-PGS are associated with their respective primary phenotypes within the CALM cohort

We first assessed the associations between ADHD and Intelligence PGS with their respective behavioural and cognitive primary phenotypes within the transdiagnostic CALM cohort. We found that ADHD-PGS were positively associated with Hyperactivity/Impulsivity scores (r^2^ = 0.03; Beta=0.17 SE=0.04; p=3.52e-05) and Inattention (r^2^ = 0.02; Beta=0.14 SE=0.04; p=1.57e-03) (**Fig. 1, Table S1**). These associations remained significant after covariation with Index of Multiple Deprivation (IMD, **Table S2**). Sensitivity analysis removing ADHD-diagnosed participants showed that the associations between ADHD-PGS and ADHD traits were not solely observed within or biased by the presence of participants with this diagnosis (**Table S3**).

**Fig. 1.**
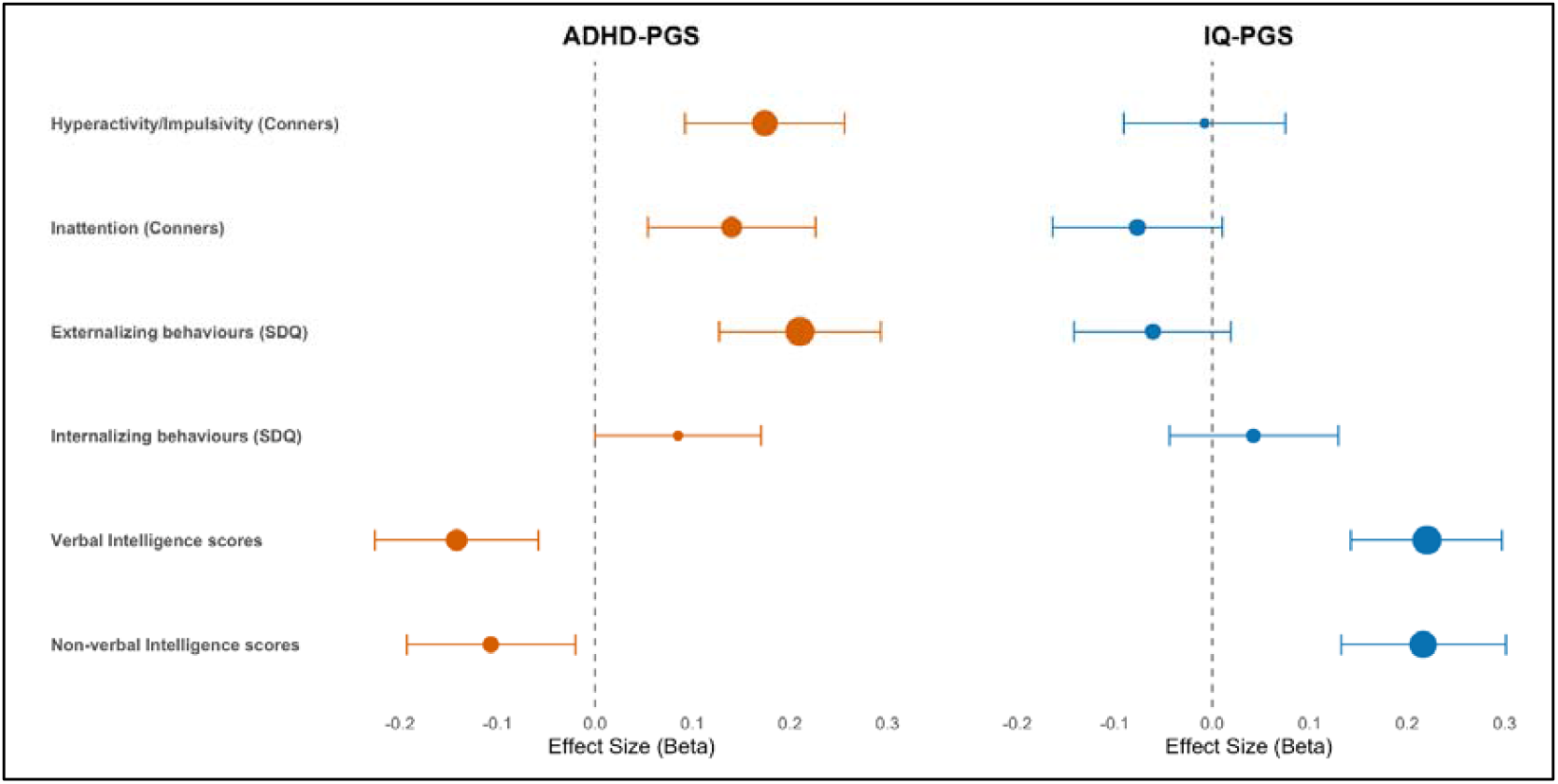
ADHD-PGS and IQ-PGS associations with dimensional measures. Forest plots showing standardized effect size (Beta) and 95% CI (error bars) from the linear regressions between ADHD and Intelligence-PGS and behavioural and cognitive measures within the CALM cohort. Vignette size corresponds to the linear regression P values (Table S1)

Intelligence-PGS were positively associated with verbal (r^2^ = 0.05; Beta=0.22 SE=0.04; p=4.00e-08) and non-verbal (r^2^ = 0.05; Beta=0.22 SE=0.04; p=7.41e-07) intelligence test scores (**Fig. 1, Table S1**), and these associations remained significant after covariation with IMD (**Table S2**).

### ADHD-PGS are associated with behavioural and cognitive dimensions beyond ADHD-related characteristics

We then analysed whether the impact of ADHD-PGS on behavioural traits can be observed beyond the narrowly defined dimensions of inattention and hyperactivity. ADHD-PGS showed positive association with externalising behaviours encompassing SDQ Conduct and Hyperactivity/Inattention scales (r^2^= 0.04; Beta=0.21 SE 0.04; p=1.06e-06), but not with internalising behaviours encompassing Emotional and Peer problems scales (r^2^= 0.01; Beta=0.09 SE=0.04; p=5.19e-02) (**Fig. 1, Table S1**). With respect to cognitive dimensions, ADHD-PGS showed negative associated with verbal (r^2^= 0.02; Beta=-0.14 SE=0.04; p=1.02e-03), and non-verbal (r^2^= 0.01; Beta=-0.11 SE=0.04; p=1.57e-02) intelligence (**Fig 1, Table S1**). All these associations remained significant after covariation with IMD (**Table S2**) or removing participants diagnosed for ADHD (**Table S3**).

### Intelligence-PGS show no association with behavioural dimensions beyond IQ

Intelligence-PGS were not significantly associated with ADHD inattention or hyperactivity scores, neither with externalising or internalising behavioural scores (**Fig. 1** and **Tables S1**; see **Table S2** for values covarying with IMD). Statistical power estimated from predicted variance explained (see **Materials and Methods**) were similar for both ADHD and Intelligence PGS, with slightly higher values for Intelligence-PGS (5.06% for ADHD-PGS and 5.11% for Intelligence-PGS, **Table S6**), confirming that the lack of association between Intelligence-PGS and behavioural dimensions was not due to reduced statistical power specifically for the intelligence-PGS.

### ADHD-PGS contributes to hierarchical factors of mental health within the CALM cohort

To assess whether ADHD and Intelligence PGS were associated with mental health, considered as a hierarchical tiered set of inter-related factors, we conducted regression analysis using the HiTOP factor scores calculated for CALM participants with data availability (see **Materials and Methods** for details). ADHD-PGS were positively associated with the level 1 general factor of psychopathology (r^2^ = 0.03; Beta=0.17 SE=0.07; p=1.37e-02), the level 2 broad externalizing factor (r^2^ = 0.03; Beta=0.17 SE=0.07; p=1.06e-02) and the level 3 social maladjustment factor (r^2^ = 0.03; Beta=0.19 SE=0.07; p=6.42e-03) (**Fig. 2, Table S4**). However, these associations were no longer significant when covarying with IMD (**Table S5**).

**Fig. 2.**
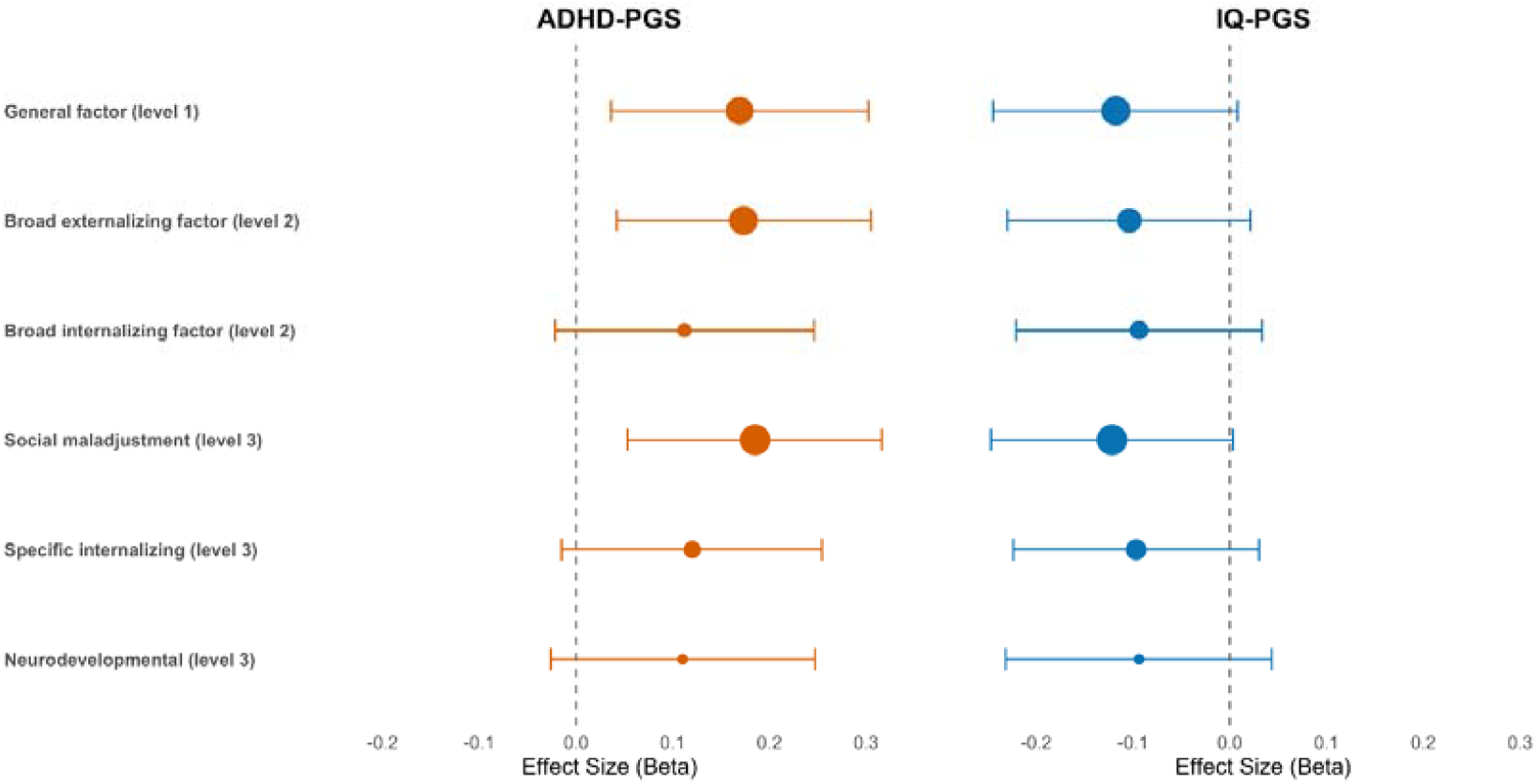
ADHD-PGS and IQ-PGS associations with HiTOP factor scores. Forest plots showing standardized effect size (Beta) and 95% CI (error bars) from the linear regressions between ADHD and Intelligence-PGS and psychopathology factors (HiTOP) within the CALM cohort. Vignette size corresponds to the linear regression P values (Table S4)

Intelligence-PGS showed no significant association with any of the HiTOP psychopathology factors, with or without IMD as a covariate (**Fig. 2, Table S4 and S5**).

Consistent with a smaller sample size for the PGS-HiTOP scores association analyses, statistical power estimates were slightly reduced (5.01% for ADHD-PGS and 5.02% for Intelligence-PGS, **Table S6**). Again, power values were similar for both PGS, thus, the lack of associations between the intelligence-PGS and HiTOP scores cannot be explained by a reduction in power specifically for intelligence-PGS.

## Discussion

We examined polygenic contributions to behavioural and cognitive variation in the CALM cohort, a clinically referred paediatric sample characterized by high levels of neurodevelopmental heterogeneity, comorbidity and learning difficulties. This transdiagnostic design provides an opportunity to test the generalizability of well-established polygenic associations for ADHD and intelligence in a setting that more closely reflects the complexity encountered in clinical practice, where diagnostic boundaries are often blurred and multiple domains of functioning are affected. Using summary statistics from the largest available GWAS at the time of analysis, we derived polygenic scores for ADHD and intelligence and evaluated their associations not only with corresponding primary phenotypes (ADHD symptoms and IQ), but also with broader behavioural dimensions assessed by the SDQ and with latent psychopathology factors derived from a hierarchical model of psychopathology (HiTOP). Our approach moves beyond single-phenotype prediction and directly examines the specificity and overlap of polygenic influences across multiple levels of phenotypic organization within a complex, transdiagnostic clinical sample.

First, we found that ADHD-PGS were associated with hyperactivity-impulsivity and inattention questionnaire scores, and Intelligence-PGS with verbal and non-verbal intelligence test scores. This confirms that GWAS-derived metrics of common genomic variation are relevant to their expected dimensions of neurodevelopmental variation despite the inclusive ascertainment criteria, high levels of co-occurring diagnoses, and sub-diagnostic difficulties of the CALM cohort. Enrichment for multi-dimensional behavioural and cognitive difficulties within the cohort means that PGS-trait relationships can be observed despite a relatively small sample size. Although a high proportion of the cohort had received a diagnosis of ADHD from community services prior to recruitment to CALM, removing these participants from analyses did not alter results. These results provide a starting point for future explorations of the associations between PGS, cognitive functions and neuroimaging correlates, and longitudinal associations within the CALM cohort.

Second, we found that PGS derived from ADHD meta-GWAS statistics were associated with neurodevelopmental dimensions beyond the core ADHD diagnostic symptoms, extending into externalising difficulties and verbal intelligence. This is consistent with previous studies that have found that higher ADHD-PGS are also associated with elevated neurodevelopmental, externalizing, and depressive and anxiety symptoms in diagnostic [47] and population studies [12, 48]. There are possible explanations for these results. The SDQ externalising score incorporates a hyperactivity subscale, so the association between ADHD-PGS and externalising scores may be driven by this subscale. The relationship between ADHD and language/verbal difficulties is well-documented both in CALM and in the broader literature [49, 50]. A single dimension of behavioural difficulties that includes both hyperactivity and inattention, was strongly and negatively associated with pragmatic communication skills within the CALM cohort [50]. The case groups contributing to ADHD-GWAS studies from which the ADHD-PGS is drawn likely included many individuals with ADHD plus additional behavioural symptoms plus reduced verbal intelligence, meaning that the ADHD-PGS may summate genomic variation contributing to each of these separate dimensions [5]. The symptoms assessed by ADHD questionnaires (particularly for the inattention subscale) are diverse and incorporate cognitive and behavioural features, meaning cross-over associations of the ADHD-PGS with diverse neurodevelopmental dimensions. Alternatively, genomic variation contributing to risk of ADHD symptoms may also contribute to risk of these additional phenotypes (a truly transdiagnostic genetic effect), evidenced by the significant genetic correlations between ADHD risk variants and IQ and educational attainment [11, 14]. Future studies could test these alternative explanations.

With respect to internalizing symptoms, we observed a weak positive association with ADHD-PGS that did not reach statistical significance. In contrast, Schlag et al. (2022) reported a robust association between ADHD-PGS and peer problems, a specific subscale of internalizing symptoms construct measured using the SDQ [51]. Their analyses were conducted in two large UK population-based cohorts (ALSPAC, N = 6,174; TEDS, N = 7,112), providing substantially greater power to detect polygenic effects than in our sample. Although non-significant, the direction of association in our study was consistent with their findings. Importantly, the CALM cohort differs from these population-based samples in that it is transdiagnostic and enriched for children with learning difficulties, which may introduce greater phenotypic heterogeneity. When participants with an ADHD diagnosis were excluded—thereby creating a sample that more closely resembles a general population-based study with low rate of clinical symptoms—the association between ADHD-PGS and internalizing symptoms reached statistical significance despite the reduced sample size (**Table S3**). Taken together these findings highlight that differences in sample composition, rather than power alone, may modulate detectability of genetic and behavioural associations, and suggest that ADHD-PGS contribution to internalizing behaviours may be sample dependent, reflecting complex influences that require further investigation.

Contrary to our transdiagnostic effects showed by the ADHD-PGS, Intelligence-PGS showed a relationship only with verbal and non-verbal IQ test scores and did not cross-associate with any behavioural measures within the CALM sample. Several features of the CALM sample may contribute to this pattern. The cohort is clinically referred and enriched for learning difficulties, with high comorbidity and potentially restricted variation in cognitive outcomes, which may limit the detectability of cross-trait associations in a modestly sized sample. Whilst ADHD-PGS have been shown to correlate with intelligence and educational attainment in population-based cohorts [11, 14], the reverse relationship between cognitive abilities and mental health seems to be strongly influenced by environmental factors [52]. Additionally, cross-trait prediction in deeply phenotyped but smaller cohorts inherently yield smaller effect sizes compared with within-trait prediction in large GWAS. However, the lack of intelligence-PGS and ADHD-traits associations cannot be explained by a reduction in intelligence-PGS statistical power, as both PGS showed similar power values when estimated based on predicted variance explained using the R package AVENGEME [46] (Suppl **Table S6**). Although the power estimates indicated limited ability to detect the expected polygenic signal in our target sample, this reflects conservative assumptions derived from GWAS architecture and same-trait prediction. Our analyses instead focused on cross-trait, dimensional associations in a deeply phenotyped cohort, with an observed variance explained within the range reported for dimensional ADHD symptom scores (∼0.7–3.3%) [53]. Finally, the observed pattern of effects— positive associations with externalizing and inattention measures and negative associations with cognitive traits—was directionally consistent with known genetic correlations of ADHD [14], supporting the interpretation of partially separable polygenic contributions to cognitive and behavioural domains.

Using the transdiagnostic CALM cohort, we tested whether ADHD polygenic risk is associated with general psychopathology, more specific externalizing or neurodevelopmental dimensions, or across multiple hierarchical levels in childhood. In addition, by examining ADHD-PGS alongside Intelligence-PGS within the same HiTOP framework, we directly compared the breadth and specificity of behavioural and cognitive polygenic effects, extending prior work beyond single-liability analyses. Here again, we found that Intelligence-PGS may not have predictive value in explaining psychopathological variation within the sample. These findings seem not to be caused by a reduction in power specifically for intelligence-PGS, as power values were similar for both ADHD and Intelligence-PGS (**Table S6**). ADHD-PGS analyses in contrast, suggest that there are polygenic contributions to an overarching p factor within CALM, indexing general susceptibility to emotional-behavioural symptoms as has been shown previously [12, 24]. Interestingly, we did not detect any association between ADHD-PGS and the lower-level “neurodevelopmental” factor which encompasses inattention, hyperactivity/impulsivity and executive function. Instead, ADHD-PGS were associated with “social maladjustment”, encompassing conduct-related and peer-related problems assessed by the SDQ. We speculate that this observed association may be explained by the high comorbidity between ADHD and conduct disorders [2]. A within-case analysis of individuals with comorbid ADHD and conduct disorders showed a positive linear relationship between ADHD-PGS and conduct problems, especially aggression [54]. Alternatively, the relationship between ADHD-PGS and verbal abilities, rather than a direct effect on behavioural characteristics, may have also contributed to these observations. The importance of communication abilities (pragmatic skills in particular) and their relationship with behavioural symptomatology have been emphasised in previous CALM analyses [50], and future hypothesis-driven studies could investigate the links between ADHD-PGS and language development. However, it is important to note that the sample available for PGS-HiTop analysis was smaller, with the concomitant reduction in power, thus, the specificity of associations should be treated with caution.

We found that the PGS associations with cognitive and behavioural measures were insensitive to socioeconomic status approximated using IMD. These results perhaps reflect the limited range of IMD within the CALM sample (Figure S1), with an overage above the national median, and are consistent with previous work conducted in large cohort showing that genetic influence on educational and cognitive outcomes is larger within more socioeconomically deprived contexts [55]. Associations between ADHD-PGS and HiTop psychopathology factors were weakened (or rendered non-significant) when co-varying for IMD. This may indicate that the observed associations are artefacts of co-variation, or that the sample is under-powered to detect significant associations once co-variates are accounted for, or alternatively that there are important gene x environment interactions at play. Although strongly influential, SES is only one of the environmental factors that contribute to behavioural and environmental outcomes [37]. Future studies could explore gene x environment interactions within this sample by assessing the impact of other important environmental factors, such as parental education and perinatal health, highly relevant in neurodevelopmental outcomes [56].

In summary, we have investigated a unique transdiagnostic sample and found that PGS contribute significantly and specifically to behavioural characteristics and cognitive abilities. This is a first step toward investigating links between genomic variation, brain development and cognitive processes contributing to learning and behavioural problems within transdiagnostic populations.

## Acknowledgments

Genotyping was carried out within the “Healthy minds 0-100 years: Optimising the use of European brain imaging cohorts (Lifebrain)” project, funded by EU Horizon2020 (Grant agreement number: 732592). We are grateful to Professor Sadaf Farooqi and her laboratory for DNA extractions and storage, and to NIHR BRC-MRC BioRepository for sample archiving.

The Centre for Attention Learning and Memory (CALM) research clinic was based at the MRC Cognition and Brain Sciences Unit, University of Cambridge, funded by the MRC through UKRI (https://calm.mrc-cbu.cam.ac.uk/). The authors wish to thank the many professionals working in children’s services in the South-East and East of England for their support, and the children and their families for contributing to the project. The lead investigators are Duncan Astle, Kate Baker, Susan Gathercole, Joni Holmes, Rogier Kievit and Tom Manly. Data collection is assisted by a team of researchers and PhD students that includes Danyal Akarca, Joe Bathelt, Marc Bennett, Madalena Bettencourt, Giacomo Bignardi, Sarah Bishop, Erica Bottacin, Lara Bridge, Diandra Brkic, Annie Bryant, Sally Butterfield, Elizabeth Byrne, Gemma Crickmore, Edwin Dalmaijer, Fánchea Daly, Tina Emery, Laura Forde, Grace Franckel, Delia Furhmann, Andrew Gadie, Sara Gharooni, Jacalyn Guy, Erin Hawkins, Rebeca Ianov-Vitanov, Christian Iordanov, Agnieszka Jaroslawska, Sara Joeghan, Amy Johnson, Jonathan Jones, Silvana Mareva, Jessica Martin, Elise Ng-Cordell, Sinead O’Brien, Cliodhna O’Leary, Joseph Rennie, Andrea M. Santangelo, Ivan Simpson-Kent, Roma Siugzdaite, Tess Smith, Stepheni Uh, Maria Vedechkina, Francesca Woolgar, Natalia Zdorovtsova, Mengya Zhang.

## Statements and Declarations

### Data Availability

Behavioural and cognitive data are available online through the CALM Managed Open Access repository: https://portal.ide-cam.org.uk/overview/1158. Genetic data are available via collaboration with CALM.

### Ethics

All procedures complied with the ethical standards of the national and institutional committees on human experimentation and with the Helsinki Declaration of 1975, as revised in 2008. All procedures involving human participants were approved by the National Health Service, Health Research Authority NRES Committee East of England (REC: 13/EE/0157, IRAS 127675).

### Consent to participate

Informed consent was obtained from legal guardians.

### Authors’ contributions

Conceptualization (KB, DA, DB, AS); Formal analysis (AS); Funding acquisition (KB); Investigation (JH, DA, SM, CALM team); Resources (OO, LB); Writing – original draft (AS, KB); Writing – review & editing (all authors).

