## Supplementary material for "ADHD and intelligence polygenic scores associations with developmental dimensions in children with attention, learning and memory difficulties"

### **Contents**

Supporting materials and methods

Supporting Figure S1

Supporting Figure S2

Supporting Tables S1-S6

### Supporting materials and methods

***Genotyping, QC filtering and imputation procedures***

DNA samples were collected from saliva in vials using the Oragene® DNA self-collection kits (DNA Genotek Inc). Children were asked to produce a saliva sample by first rubbing their cheeks gently for 30 s to create saliva, and then they were asked to spit in a pot. For children who find it hard to create saliva, a small amount (max ¼ tsp) of white table sugar was available to place on the child’s tongue. The saliva samples were stored in the Oragene® kits at room temperature (15–30 °C) until DNA extraction, which was performed using prepIT∙L2P Kit (DNA Genotek Inc) following manufacturer instructions. Once extracted, DNA was stored at -80 °C at the Wellcome Trust-MRC Institute of Metabolic Science at Addenbrooke’s Hospital and subsequently transferred to the BioRepository for long-term storage.

Genome-wide SNP genotyping was performed using the Global Screening Array with shared custom-content (GSA, Illumina, Inc., USA) on an iScan instrument according to manufacturer’s recommendations. The raw intensity data was subjected to genotype calling, extensive pre-imputation quality control and SNP imputation using an automated computational workflow described previously (Hong et al., 2020, 2021). A total of 7 samples were excluded because of low quality DNA or evidence of cross-contamination. We estimated the genetic sex of participants using the command --check-sex 0.25 0.75 in PLINK (Purcell et al., 2007). This led to the exclusion of 10 participant in whom genetic sex could not be determined unambiguously or in whom there was a discrepancy with the clinically reported sex. One more sample was removed because of excessive missing data (--mind 0.05). The LD pruned dataset was also used for principal component analysis (PCA; using PLINK command ‘--pca') along with the reference dataset of the 1000 Genome Project Consortium Phase 3 (The 1000 Genomes Project Consortium, 2015) to assign ancestry using the five 1000G super-populations by k-nearest neighbour classification (k-NN; k=9; using R package ‘class’ in R 2.3.2 (Venables & Ripley, 2002). Only European-descent samples were used in the subsequent analyses, 40 non-European samples were removed.

Genotype imputation was performed on the QC’ed and filtered data with MiniMac3 (Das et al., 2016) software using the "Haplotype Reference Consortium" (HRC; v1.1 [EGAD00001002729 including 39,131,578 SNPs from ~11K individuals]) reference panel (McCarthy et al., 2016). Overall, this procedure resulted in 39,131,578 imputed SNPs.

***PGS calculation***

To aggregate data on multiple variants per individual we computed PGS for each individual as described previously (Choi et al., 2020). Allele status and effect-size estimates were taken from the summary statistics of the largest ADHD (Demontis et al., 2023) and Intelligence (Savage et al., 2018) meta-GWAS published to date. First, we removed duplicate and ambiguous SNPs (A/T and C/G) from the list of considered variants. We only used SNPs with MAF >0.05 (because our target dataset is N<1000 samples), and imputation quality r^2^ >0.8. The remaining variants were 5,685,483 (out of 6,774,224 SNPs) for ADHD-GWAS, and 4,673,194 (out of 9,295,118 SNPs) for Intelligence-GWAS. The target CALM sample did not overlap with any of these GWAS datasets. Both GWAS and the CALM genotyping data were annotated on the GRCh37/hg19 genome build. The non-referred individuals were removed, remaining only 557 samples. Further QC included filtering for samples with extreme heterozygosity (more than 3 SD from the mean), by performing first pruning (--indep-pairwise 200 50 0.25) and then calculating the F coefficient for each individual (--het), leading to the exclusion of 9 samples. Finally, 24 closely related individuals were also removed (--rel-cutoff 0.125), giving a final sample size of **N=524**. Duplicate SNPs were removed using --exclude in PLINK, with 38,913,048 out of 39,131,578 variants remaining. After further removing all SNPs with MAF<0.01, P value for HWE < 1e-6, and genotyping efficiency <99% (--geno 0.01), 7,568,187 variants remained. Variant matching and recoding between the ADHD/Intelligence GWAS base and CALM target datasets was performed using R (version 3.5.3), resulting in 5,583,323 and 3,994,342 overlapping SNPs for ADHD and Intelligence, respectively.

To account for LD, we performed clumping in PLINK (--clump-p1 1, --clump-r2 0.1, --clump-kb 250) to remove the SNPs highly linked to the index SNP and with low association to the GWAS-specific phenotype. To account for population stratification, we performed a PCA on the pruned SNP pool (--indep-pairwise 200 50 0.25) and extracted the first six PC (--pca 6). Actual PGS were computed using the --score command for a variety of P-value thresholds (P_t_) in the primary GWAS data (0.001, 0.05, 0.10, 0.20, 0.30, 0.40, 0.50).

### Supporting Figure S1

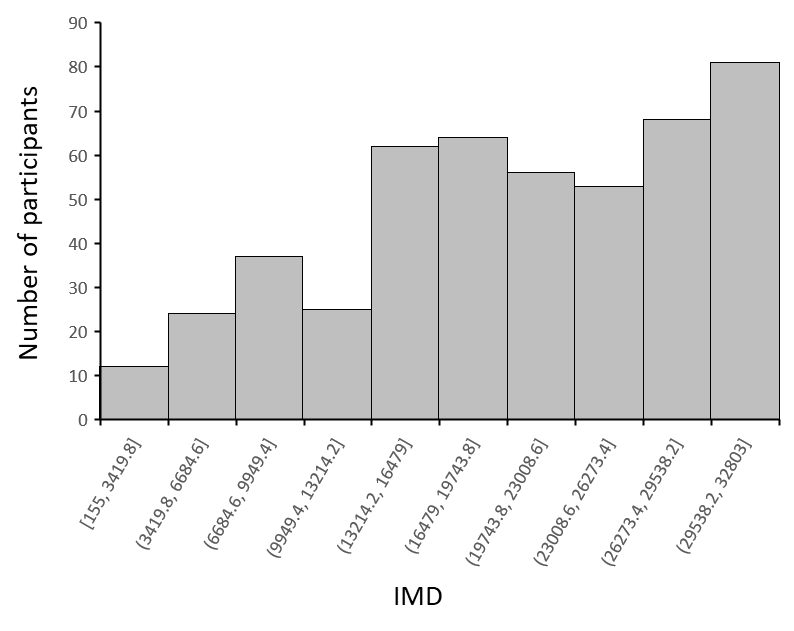

**Figure S1**: Frequency distribution of the Index of Multiple Deprivation (IMD) in deciles.

### Supporting Figure S2

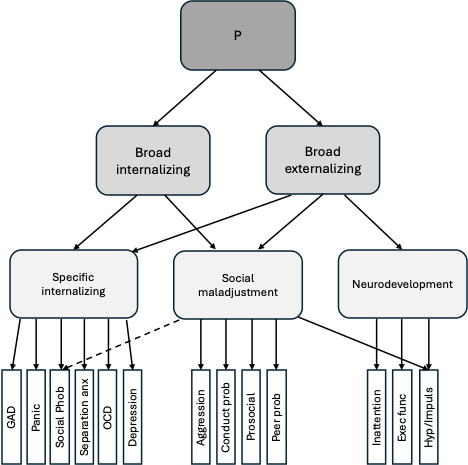

**Figure S2**: Hierarchical Structure of Psychopathology in the Centre for Attention Learning and Memory (CALM) Sample, from Holmes et al. (2021). Dashed lines represent negative associations. For simplicity, correlation values were omitted. GAD: General Anxiety Disorder; OCD: Obsessive Compulsive Disorder; Panic: Panic disorder; Social phob: Social phobia; Separation anx: Separation anxiety (all from RCADS); Conduct prob; Conduct problems; Prosocial: Prosocial behavior (reverse coded; SDQ); Peer prob: Peer relations; Exec func: Executive functions; Hyp/Impuls: Hyperactivity and impulsivity (all from CPSF). P: general psychopathology.

### Supporting Tables

| **Table S1: Linear regression between PGS and behavioural and cognitive measures with sex, age and first 6PC as covariates, N=524 (* p< Benjamini-Hochberg critical value).** | | | | | | | |
| --- | --- | --- | --- | --- | --- | --- | --- |
| **Measures** | **P t** | **R2** | **BETA** | **SE** | **P value** | **Rank** | **BH critical value** |
| **ADHD-PGS** |  |  |  |  |  |  |  |
| Extern* | 0.200 | 0.043 | 0.210 | 0.043 | 1.06E-06 | 1 | 0.008 |
| Hyp/Imp* | 0.400 | 0.030 | 0.174 | 0.042 | 3.52E-05 | 2 | 0.017 |
| Verbal Intel* | 0.300 | 0.020 | -0.142 | 0.043 | 1.02E-03 | 3 | 0.025 |
| Inatt* | 0.300 | 0.019 | 0.140 | 0.044 | 1.57E-03 | 4 | 0.033 |
| Non-verb Intel* | 0.400 | 0.011 | -0.107 | 0.044 | 1.57E-02 | 5 | 0.042 |
| Intern | 0.001 | 0.007 | 0.085 | 0.044 | 5.19E-02 | 6 | 0.050 |
| **Intelligence-PGS** |  |  |  |  |  |  |  |
| Verbal Intel* | 0.001 | 0.054 | 0.220 | 0.040 | 4.00E-08 | 1 | 0.008 |
| Non-verb Intel* | 0.500 | 0.046 | 0.216 | 0.043 | 7.41E-07 | 2 | 0.017 |
| Inatt | 0.500 | 0.006 | -0.077 | 0.044 | 8.24E-02 | 3 | 0.025 |
| Extern | 0.001 | 0.004 | -0.061 | 0.041 | 1.36E-01 | 4 | 0.033 |
| Intern | 0.050 | 0.002 | 0.042 | 0.044 | 3.36E-01 | 5 | 0.042 |
| Hyp/Imp | 0.100 | 0.000 | -0.008 | 0.042 | 8.58E-01 | 6 | 0.050 |

| **Table S2: Linear regression between PGS and behavioural and cognitive measures with sex, age, first 6PC, and IMD as covariates, N=524 (* p< Benjamini-Hochberg critical value).** | | | | | | | |
| --- | --- | --- | --- | --- | --- | --- | --- |
| **Measures** | **P t** | **R2** | **BETA** | **SE** | **P value** | **Rank** | **BH critical value** |
| **ADHD-PGS** |  |  |  |  |  |  |  |
| Extern* | 0.200 | 0.041 | 0.206 | 0.044 | 3.05E-06 | 1 | 0.008 |
| Hyp/Imp* | 0.400 | 0.024 | 0.160 | 0.043 | 2.62E-04 | 2 | 0.017 |
| Inatt* | 0.400 | 0.014 | 0.118 | 0.046 | 9.90E-03 | 3 | 0.025 |
| Verbal Intel* | 0.300 | 0.011 | -0.108 | 0.045 | 1.69E-02 | 4 | 0.033 |
| Non-verb Intel* | 0.400 | 0.009 | -0.100 | 0.047 | 3.49E-02 | 5 | 0.042 |
| Intern | 0.001 | 0.005 | 0.073 | 0.044 | 1.02E-01 | 6 | 0.050 |
| **Intelligence-PGS** |  |  |  |  |  |  |  |
| Non-verb Intel* | 0.500 | 0.049 | 0.228 | 0.046 | 8.16E-07 | 1 | 0.008 |
| Verbal Intel* | 0.001 | 0.043 | 0.198 | 0.041 | 1.77E-06 | 2 | 0.017 |
| Intern | 0.050 | 0.003 | 0.060 | 0.045 | 1.86E-01 | 3 | 0.025 |
| Inatt | 0.500 | 0.003 | -0.058 | 0.045 | 2.02E-01 | 4 | 0.033 |
| Extern | 0.001 | 0.003 | -0.053 | 0.048 | 2.05E-01 | 5 | 0.042 |
| Hyp/Imp | 0.200 | 0.000 | 0.008 | 0.044 | 8.58E-01 | 6 | 0.050 |

| **Table S3: Sensitivity analysis without participants diagnosed for ADHD. Linear regression between ADHD-PGS and behavioural and cognitive measures, N=405 (* p< Benjamini-Hochberg critical value).** | | | | | | | |
| --- | --- | --- | --- | --- | --- | --- | --- |
| **Sex, age and first 6PC as covariates** | | | | | | | |
| **Measures** | **P t** | **R2** | **BETA** | **SE** | **P value** | **Rank** | **BH critical value** |
| Extern* | 0.300 | 0.027 | 0.168 | 0.049 | 7.02E-04 | 1 | 0.008 |
| Non-verb Intel* | 0.050 | 0.020 | -0.143 | 0.050 | 4.29E-03 | 2 | 0.017 |
| Verbal Intel* | 0.500 | 0.016 | -0.135 | 0.052 | 1.02E-02 | 3 | 0.025 |
| Hyp/Imp* | 0.500 | 0.014 | 0.120 | 0.048 | 1.30E-02 | 4 | 0.033 |
| Inatt* | 0.300 | 0.015 | 0.133 | 0.054 | 1.46E-02 | 5 | 0.042 |
| Intern* | 0.001 | 0.010 | 0.104 | 0.050 | 3.93E-02 | 6 | 0.050 |
| **Sex, age, first 6PC, and IMD as covariates** | | | | | | | |
| **Measures** | **P t** | **R2** | **BETA** | **SE** | **P value** | **Rank** | **BH critical value** |
| Extern* | 0.400 | 0.028 | 0.173 | 0.051 | 8.24E-04 | 1 | 0.008 |
| Non-verb Intel* | 0.050 | 0.017 | -0.139 | 0.053 | 9.68E-03 | 2 | 0.017 |
| Hyp/Imp* | 0.500 | 0.012 | 0.116 | 0.050 | 2.13E-02 | 3 | 0.025 |
| Inatt | 0.400 | 0.012 | 0.120 | 0.057 | 3.61E-02 | 4 | 0.033 |
| Verbal Intel | 0.300 | 0.010 | -0.108 | 0.055 | 4.90E-02 | 5 | 0.042 |
| Intern | 0. 001 | 0.009 | 0.096 | 0.052 | 6.31E-02 | 6 | 0.050 |

| **Table S4: Linear regression between PGS and HiTOP factors with sex, age and first 6PC as covariates, N=218 (* p< Benjamini-Hochberg critical value).** | | | | | | | |
| --- | --- | --- | --- | --- | --- | --- | --- |
| **Factors** | **P t** | **R2** | **BETA** | **SE** | **P value** | **Rank** | **BH critical value** |
| **ADHD-PGS** |  |  |  |  |  |  |  |
| Social adj* | 0.050 | 0.033 | 0.185 | 0.067 | 6.42E-03 | 1 | 0.008 |
| Broad Ext* | 0.050 | 0.029 | 0.173 | 0.067 | 1.06E-02 | 2 | 0.017 |
| P factor* | 0.050 | 0.028 | 0.169 | 0.068 | 1.37E-02 | 3 | 0.025 |
| Specific Int | 0.050 | 0.014 | 0.120 | 0.069 | 8.23E-02 | 4 | 0.033 |
| Broad Int | 0.050 | 0.012 | 0.112 | 0.069 | 1.03E-01 | 5 | 0.042 |
| Neurodev | 0.400 | 0.012 | 0.111 | 0.070 | 1.14E-01 | 6 | 0.050 |
| **Intelligence-PGS** |  |  |  |  |  |  |  |
| Social adj | 0.001 | 0.016 | -0.122 | 0.064 | 5.80E-02 | 1 | 0.008 |
| P factor | 0.001 | 0.015 | -0.118 | 0.064 | 6.74E-02 | 2 | 0.017 |
| Broad Ext | 0.001 | 0.012 | -0.104 | 0.064 | 1.04E-01 | 3 | 0.025 |
| Specific Int | 0.001 | 0.010 | -0.097 | 0.065 | 1.36E-01 | 4 | 0.033 |
| Broad Int | 0.001 | 0.010 | -0.094 | 0.065 | 1.49E-01 | 5 | 0.042 |
| Neurodev | 0.500 | 0.008 | -0.094 | 0.070 | 1.81E-01 | 6 | 0.050 |

| **Table S5: Linear regression between PGS and HiTOP factors with sex, age, first 6PC, and IMD as covariates, N=218 (* p< Benjamini-Hochberg critical value).** | | | | | | | |
| --- | --- | --- | --- | --- | --- | --- | --- |
| **Factors** | **P t** | **R2** | **BETA** | **SE** | **P value** | **Rank** | **BH critical value** |
| **ADHD-PGS** |  |  |  |  |  |  |  |
| Social adj | 0.050 | 0.018 | 0.137 | 0.066 | 3.92E-02 | 1 | 0.008 |
| Broad Ext | 0.050 | 0.017 | 0.133 | 0.066 | 4.47E-02 | 2 | 0.017 |
| P factor | 0.050 | 0.013 | 0.112 | 0.066 | 9.14E-02 | 3 | 0.025 |
| Neurodev | 0.400 | 0.009 | 0.097 | 0.072 | 1.77E-01 | 4 | 0.033 |
| Specific Int | 0.050 | 0.004 | 0.065 | 0.068 | 3.36E-01 | 5 | 0.042 |
| Broad Int | 0.050 | 0.004 | 0.059 | 0.068 | 3.90E-01 | 6 | 0.050 |
| **Intelligence-PGS** |  |  |  |  |  |  |  |
| Neurodev | 0.500 | 0.007 | -0.085 | 0.072 | 2.41E-01 | 1 | 0.008 |
| Social adj | 0.001 | 0.006 | -0.072 | 0.062 | 2.51E-01 | 2 | 0.017 |
| P factor | 0.001 | 0.004 | -0.062 | 0.062 | 3.19E-01 | 3 | 0.025 |
| Broad Ext | 0.001 | 0.004 | -0.059 | 0.062 | 3.43E-01 | 4 | 0.033 |
| Specific Int | 0.001 | 0.003 | -0.047 | 0.064 | 4.57E-01 | 5 | 0.042 |
| Broad Int | 0.001 | 0.002 | -0.046 | 0.064 | 4.76E-01 | 6 | 0.050 |

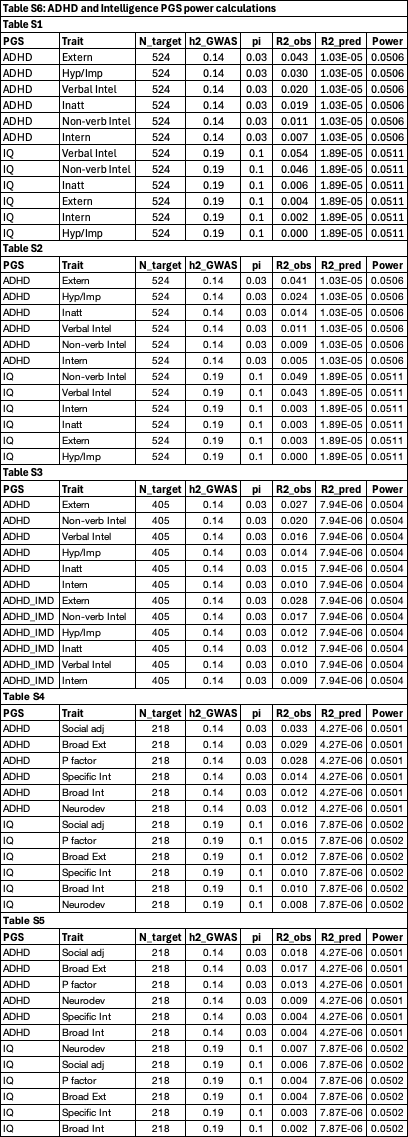
